# Testing structural interpretations beyond goodness-of-fit in a phenomenological epidemic model: A reproducibility-oriented case study

**DOI:** 10.64898/2026.10.13.26365491

**Authors:** Yoshitaka Umeno

**Affiliations:** Department of Fundamental Engineering, Institute of Industrial Science, The University of Tokyo, Tokyo, Japan

**Keywords:** epidemic modelling, phenomenological models, reproducibility, counterfactual inference, model flexibility

## Abstract

Simulation models are widely used to support counterfactual inference in settings where direct experimental validation is difficult or infeasible. However, for phenomenological models calibrated to observational time series, goodness-of-fit does not necessarily establish that the structural assumptions underlying such inferences are uniquely supported by the data. Here, we examine this issue using a published COVID-19 transmission model for Japan that incorporates empirical inputs including human mobility, variant prevalence, and vaccination and has been used to estimate the effects of alternative vaccination scenarios. Using the publicly archived code together with a reconstructed substitute for a restricted-access input file required by the released implementation, we first reproduced the published model behaviour. We then performed two complementary computational stress tests. First, the vaccination-related input was suppressed and the remaining parameters were re-estimated against the original epidemic trajectory. Second, as a complementary test of trajectory-level flexibility, the model was refitted to synthetic target series with qualitatively different temporal structures while retaining the empirical mobility and variant inputs. The model retained substantial fitting capacity in both tests: suppression of the vaccination-related input caused only a limited deterioration in fit to the original trajectory, while qualitatively different synthetic trajectories could also be reproduced through parameter adjustment. Variation in fitted parameters across these conditions was also consistent with compensatory parameter behaviour. These results do not imply that vaccination had no effect or that the original model is invalid. Rather, they provide empirical evidence that, in this case, goodness-of-fit to the observed epidemic trajectory alone does not uniquely support the structural interpretation required for vaccination-related counterfactual inference. The study illustrates the value of reproducible computational stress testing as a complement to, rather than a substitute for, formal identifiability or model-comparison analyses.

## 1. Introduction

Simulation models are widely used across scientific and engineering disciplines to explore complex systems when direct experimentation is difficult, costly, or ethically constrained. In many applications, phenomenological models are calibrated to observational time series through parameter fitting while incorporating simplified representations of the processes thought to generate the observed dynamics. Such models can be useful for description, forecasting, and scenario analysis. However, close agreement between simulated and observed trajectories does not necessarily establish that the structural assumptions embedded in the model are uniquely supported by the available data. This distinction becomes particularly important when fitted models are used not only to reproduce observations, but also to interpret underlying processes or to estimate outcomes under counterfactual scenarios.

Epidemic modelling provides a prominent example. During the COVID-19 pandemic, models incorporating epidemiological structure together with empirical inputs such as human mobility, vaccination coverage, and variant prevalence were used to evaluate interventions and alternative policy scenarios [1]. In one published study, Kayano et al. developed an age-structured transmission model calibrated to national COVID-19 data from Japan and used it to estimate outcomes under alternative vaccination scenarios [2][3]. The model incorporates empirical time-series inputs for vaccination, human mobility, and variant prevalence and therefore represents a substantially more data-informed formulation than simple curve fitting. The model was used to derive quantitative estimates of infections and deaths averted under vaccination scenarios, making the interpretation of its vaccination-related component consequential for the resulting counterfactual inference. These features make the model a useful case for examining a broader methodological question: to what extent does successful reproduction of an observed epidemic trajectory constrain the structural interpretation on which a counterfactual calculation depends?

The general issues underlying this question are well recognized. Structural and practical identifiability, parameter degeneracy, structural uncertainty, and model equifinality have been extensively discussed in dynamical modelling and epidemiology [4]-[7]. The present study does not propose a formal identifiability analysis or a general model-selection framework, nor does it seek to establish these methodological concerns as new. Rather, we ask how strongly they can manifest in a concrete published model that combines multiple empirical covariates and has been used for policy-relevant counterfactual inference. Examining this question computationally in an actual published model provides information that is distinct from the general theoretical observation that different parameterizations or model structures may sometimes be compatible with the same data.

Here, we use the publicly archived implementation of the Kayano–Nishiura model. Because an input file required to execute the released code was not included in the archived package, we reconstructed the unavailable infection input data from the published results and generated a runnable input dataset. This enabled us to reproduce the published model behaviour and to conduct a reproducibility-oriented computational case study. We first establish that the published model behaviour can be approximately reproduced with the reconstructed data. We then examine whether suppressing the vaccination-related input, followed by re-estimation of the remaining parameters, necessarily causes a substantial deterioration in the fit to the original epidemic trajectory. As a complementary stress test of trajectory-level flexibility, we also re-estimate the model against synthetic target series with qualitatively different temporal structures while retaining the empirical mobility and variant inputs. Finally, we examine variation in the fitted parameters across these conditions and the sensitivity of the reproduction to assumptions used in reconstructing the age distribution. These analyses are intended to test a deliberately limited question: whether goodness-of-fit to the observed trajectory alone provides strong support for the structural interpretation required for counterfactual inference in this particular model.

## 2. Simulation model and computational stress tests

### 2.1 Reference time-series model

To make the discussion concrete, we base our analysis on a previously published age-structured time-series transmission model for COVID-19 in Japan [2]. In what follows, we refer to this as the Kayano–Nishiura model. The model extends a classical susceptible–infected–recovered (SIR)–type framework by dividing the population into ten age groups and incorporating empirical drivers such as vaccination, human mobility and variant prevalence.

The number of infected individuals in age group *a* {*a* ∈ ℕ| *a* ≤ 10} at time *t*, denoted *i*_*a*_(*t*), is expressed as

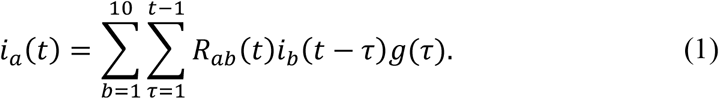

Here, *g* denotes a probability density function, which is assumed to follow a Weibull distribution. *R*_*ab*_ represents the effective reproduction number from age group *b* to *a*, and is given by,

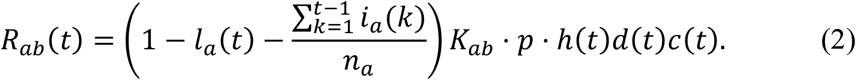

Here, *n*_*a*_ denotes the population size of age group *a*, and *l*_*a*_ is a function representing the infection suppression effect of vaccination (described later). *K*_*a b*_ is a coefficient matrix corresponding to the contact frequency between age groups *a* and *b*, and *p* is an overall scaling factor (one of the adjustable parameters of the model). The functions *h, d*, and *c* represent the temporal change of human mobility, changes in infectivity due to the Delta variant mutation, and the effects of consecutive holidays, respectively. These functions are determined based on empirical data as follows:

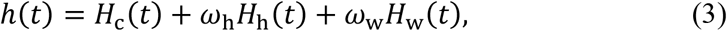

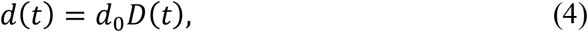

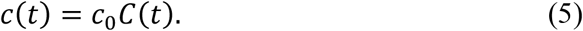

*H*_c_(*t*), *H*_h_(*t*) and *H*_w_(*t*) represent the actual time-series data of human mobility in Japan corresponding to community, household, and workplace settings, respectively, as reported in Google’s COVID-19 Community Mobility Report [8]. The parameters ω_h_ and *ω*_w_ are adjustable parameters that determine the relative contributions of household and workplace mobility, respectively, when that of community mobility is fixed at 1.0 by definition (i.e. *ω*_*c*_ = 1). *D*(*t*) denotes the actual data of the proportion of Delta variant detections relative to the total number of reported cases, and *d*_0_ is the adjustable parameter controlling its contribution. *C*(*t*) is a time-series indicator of holidays lasting three days or longer, with *c*_0_ representing the corresponding adjustable parameter for its influence.

The function *l*_*a*_ is defined as follows:

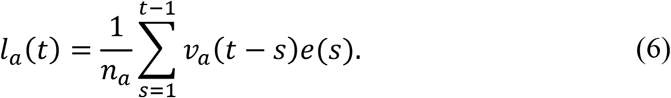

Here, *v*_*a*_(*t*) represents the vaccination rate for age group *a* on date *t*, based on actual data. The function *e*(*t*) denotes the infection suppression effect after vaccination, which gradually decreases over time following a certain period post-vaccination, reflecting the waning of vaccine efficacy.

Thus, although the Kayano-Nishiura model is a phenomenological model, it incorporates time-varying effects of vaccination, human mobility, and changes in infectivity due to viral variants. The mobility and other data are based on actual measurements, making it a substantially more data-informed formulation than simple curve fitting.

The model parameters *p, ω*_*h*_, *ω*_*w*_, *d*_0_ and *c*_0_ were fitted to the HER-SYS data representing the actual infection trends. These parameters are constants that do not vary over time. In the code, their initial values were set to 2.7, −0.5, −0.08, 1.1, and 1.2, respectively, and their search ranges were [0, 10], [−5, 5], [−5, 5], [0, 5], and [0, 5]. In the case where a parameter reached the upper or lower limit, the optimization was redone with a slightly widened search range. Optimization was performed using the BFGS method (Broyden–Fletcher–Goldfarb–Shanno algorithm), a variant of the Newton-Raphson method.

Although the model code was publicly archived, the input file File2.Hersys_infection.csv, which is explicitly called by the released Kayano–Nishiura code, was not included in the public archive because it contains HER-SYS surveillance data subject to legal restrictions on data sharing in Japan. The repository states that the file may be obtained through an application and approval procedure involving the National Institute of Infectious Diseases / Japan Institute for Health Security. Consequently, the released code cannot be executed directly using only the publicly downloadable files [3].

To enable independent reproduction without access to the restricted surveillance dataset, we reconstructed the required infection input by extracting discrete points from the infection trajectory reported in the Kayano–Nishiura paper and applying spline interpolation. The reconstructed data were then formatted as File2.Hersys_infection.csv so that they could be read directly by the original code. The resulting pseudo HER-SYS dataset is shown in Figure 1(a). Using this reconstructed input, we successfully executed the published code and approximately reproduced the reported model behaviour. The number of infected individuals in each age group was initially assumed to be one-tenth of the total number, as shown in Figure 1(b).

**Figure 1:**
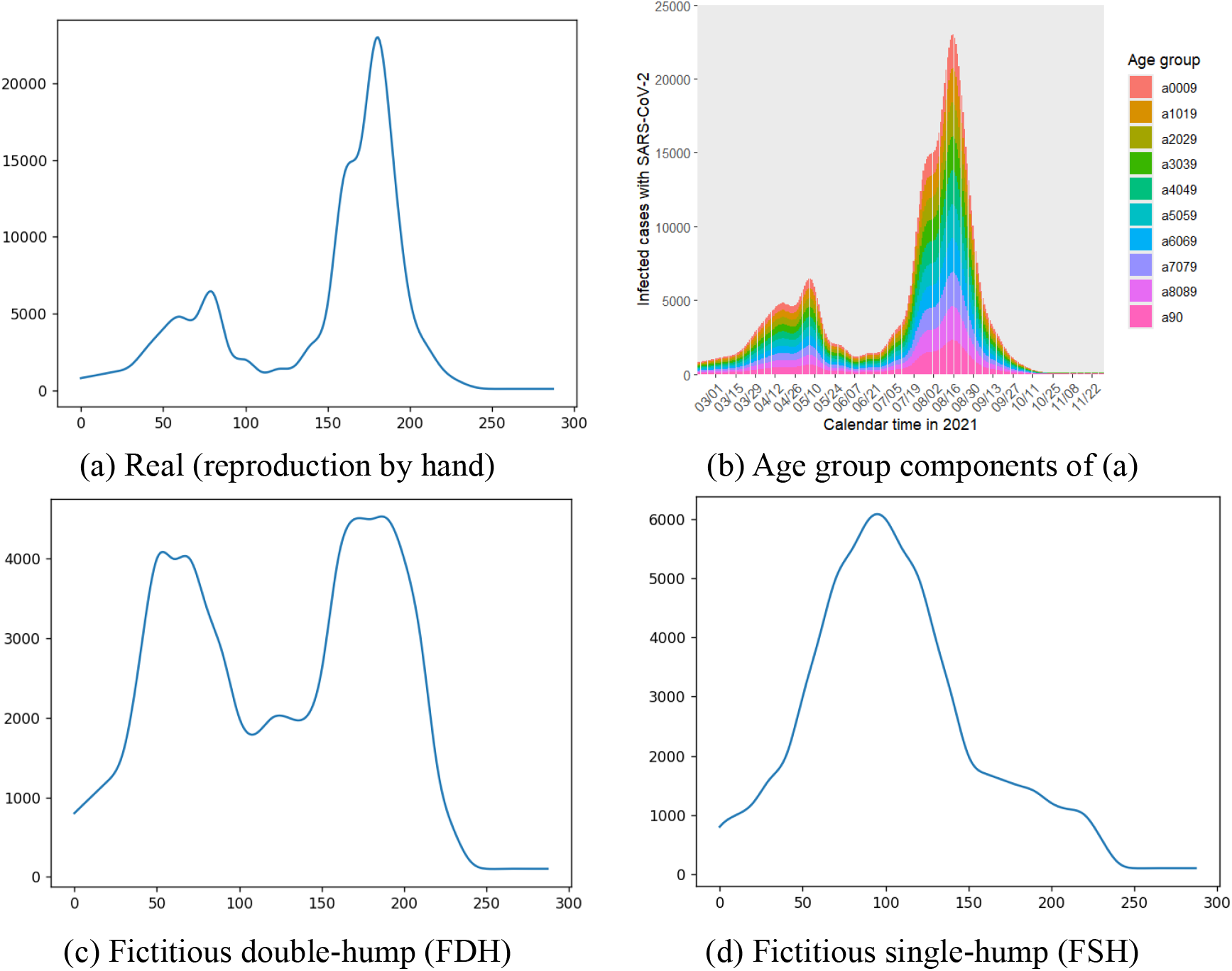
Curves of infection cases used for parameter optimization.

### 2.2 Computational stress tests

Following reproduction of the published model behaviour using the reconstructed input data, we performed two complementary computational stress tests. The primary test examined the extent to which the fit to the original epidemic trajectory depended on the vaccination-related input. A second test examined the trajectory-level flexibility of the model by fitting it to synthetic target series with qualitatively different temporal structures. These tests were designed to probe specific properties of the fitted model rather than to constitute a formal identifiability analysis or model-selection framework.

First, we suppressed the vaccination-related input by setting the number of vaccinated individuals to zero throughout the entire simulation period and re-estimated the remaining adjustable parameters against the original infection trajectory. This procedure leaves the model equations and implementation unchanged while eliminating the contribution associated with vaccination. We then examined whether parameter re-estimation could compensate for this perturbation sufficiently to retain a close fit to the reference trajectory. The purpose of this test was not to estimate the epidemiological effect of vaccination, but to assess whether goodness-of-fit to the observed trajectory strongly depends on the vaccination-related contribution within this fitted model.

Second, as a complementary stress test, we replaced the original infection curve with artificially generated synthetic target curves: a two-peaked curve with nearly equal peak heights (Fictitious Double-Hump; FDH) and a single-peaked curve (Fictitious Single-Hump; FSH) (Figure 1(c),(d)). The model parameters were then re-estimated for each target while retaining the observed time series of human mobility and Delta-variant prevalence. These experiments were not intended to establish that the model can reproduce arbitrary epidemic curves or to test structural identifiability. Rather, they were used to examine the extent to which parameter adjustment allows the model to reproduce qualitatively different temporal patterns despite retaining empirical covariates that were associated with the original epidemic trajectory.

## 3. Results

### 3.1 Reproduction of the reference epidemic trajectory

The parameters determined using the pseudo HER-SYS data are shown in Table 1 (which also includes parameters for other cases described later). The simulation results obtained with this parameter set are presented in Figure 2. It can be seen that the pseudo HER-SYS data used (also shown in the figure) are reproduced with a high accuracy. The results are highly similar to Figure 1 in the Kayano–Nishiura paper, confirming that the optimization using the pseudo HER-SYS data in this study is reliable. To complement this visual comparison, approximate normalized fit metrics were also estimated from graphically extracted trajectories of the fitted curve. For the Original case, the resulting values were RMSE/peak ≈ 0.030 and MAE/peak ≈ 0.018. Although these values are only approximate and are not intended as formal model selection criteria, they are consistent with close agreement between the fitted trajectory and the reference data.

**Table 1:** Optimized parameters for transmission model to reproduce real and fictitious curves of infected cases. Original w/o vac: original model run with the vaccination-related input set to zero. An additional derived quantity, ***p***(***ω***_***c***_ **+ *ω***_***h***_ **+ *ω***_***w***_), is included to illustrate a possible compensatory relationship between the overall transmission scaling parameter and the mobility-related contribution weights, where ***ω***_***c***_ **= 1** by definition.

| Case | $p$ | $\omega_h$ | $\omega_w$ | $d_0$ | $c_0$ | $p(\omega_c + \omega_h + \omega_w)$ |
| --- | --- | --- | --- | --- | --- | --- |
| Original | 2.468 | −0.3660 | −0.1940 | 1.047 | 1.006 | 1.09 |
| Original w/o vac | 3.092 | −0.5040 | −0.09670 | 0.7102 | 1.032 | 1.23 |
| FDH | 0.7704 | −0.04480 | 0.2151 | 1.002 | 0.9864 | 0.902 |
| FSH | 0.1240 | 5.003 | 0.4050 | 0.8710 | 0.9620 | 0.795 |

**Figure 2:**
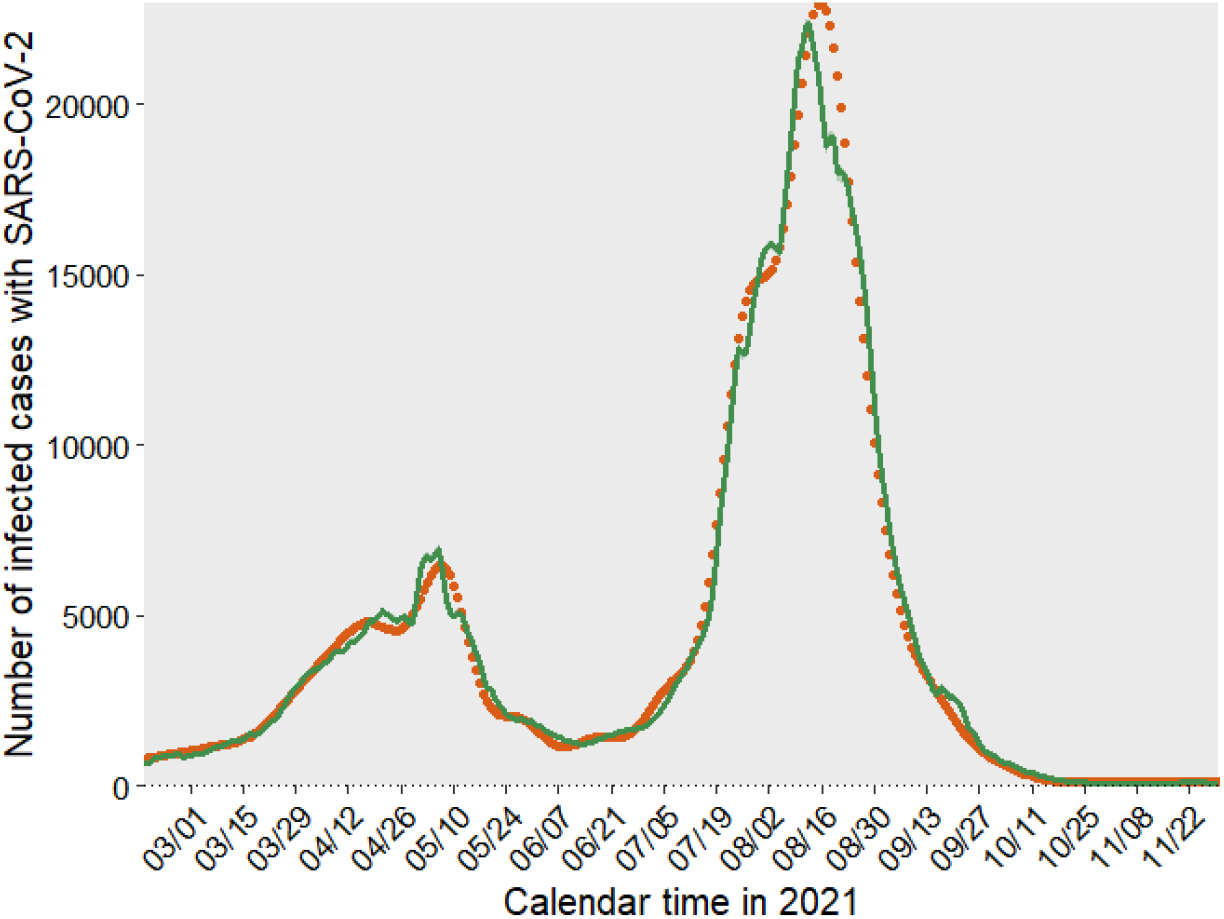
Reproduction of the reference epidemic trajectory by the Kayano–Nishiura model (solid line). Pseudo HER-SYS data are also plotted (dotted line).

Because the pseudo HER-SYS dataset required an assumed age distribution, we additionally examined the sensitivity of the Original-case fitting to this assumption. Besides the equal allocation used in the main analysis, we tested two alternative age allocations: an older-skewed allocation and a bell-shaped allocation, while preserving the same total incidence. In both cases, the fitted trajectories remained visually very similar to those obtained under the equal-allocation assumption. Approximate normalized fit errors estimated from graphically extracted trajectories were also very similar (RMSE/peak ≈ 0.031 and MAE/peak ≈ 0.019 for both alternative allocations), indicating that the main qualitative result does not depend strongly on the equal age-distribution assumption.

### 3.2 Effect of suppressing the vaccination-related input

Figure 3 shows the fitted trajectory obtained after the vaccination-related input was suppressed and the remaining adjustable parameters were re-estimated against the reference epidemic trajectory. The optimized parameters are listed in Table 1. Despite suppression of the vaccination-related contribution, the refitted model retained close agreement with the reference trajectory. Approximate normalized fit metrics estimated from graphically extracted trajectories were RMSE/peak ≈ 0.039 and MAE/peak ≈ 0.025, compared with RMSE/peak ≈ 0.030 and MAE/peak ≈ 0.018 for the Original case.

**Figure 3:**
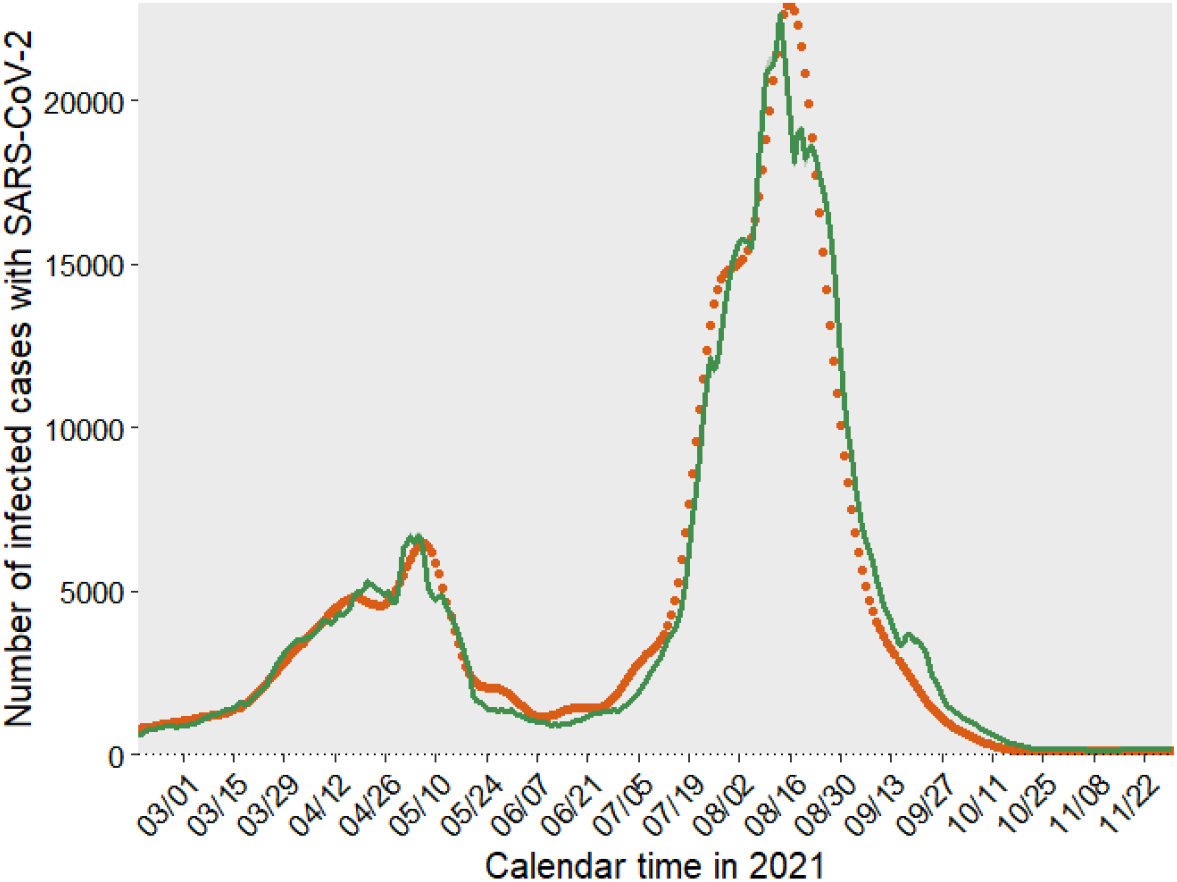
Reproduction of the reference epidemic trajectory after suppression of the vaccination-related input (solid line).

Thus, suppressing the vaccination-related input produced only a limited deterioration in trajectory-level fit after re-estimation of the remaining parameters. This result shows that close agreement with the reference epidemic trajectory can be maintained in this model even when the vaccination-related contribution is not active. It does not establish formal non-identifiability of that contribution, but demonstrates that goodness-of-fit alone does not require its inclusion to obtain a close reproduction of the reference trajectory.

### 3.3 Fitting to synthetic target trajectories

Figure 4 shows the results of fitting the Kayano–Nishiura model to the two synthetic target series. The optimized parameters are listed in Table 1. In both the FDH (Figure 4(a)) and FSH (Figure 4(b)) cases, parameter re-estimation allowed the model to reproduce the main temporal features of the target trajectories despite their qualitatively different shapes.

**Figure 4:**
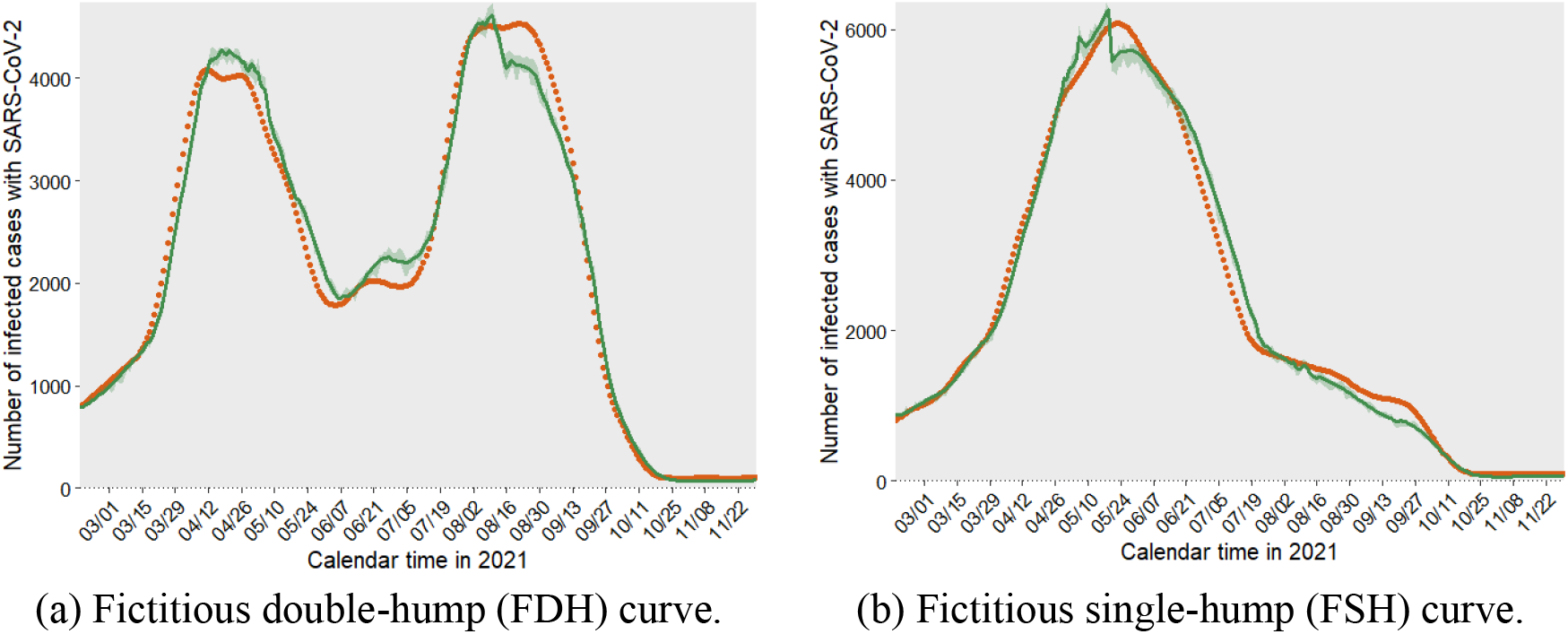
Reproduction of fictitious curves of infection cases by Kayano–Nishiura model (solid line). Artificial data used for fitting are also shown (dotted line).

Importantly, these fits continued to use the observed human mobility and Delta-variant prevalence time series. The adjustable parameters modified the relative contributions of these retained empirical drivers rather than replacing them. The results therefore show that the fitted model has substantial trajectory-level flexibility under parameter adjustment, including when the target trajectory is deliberately mismatched with empirical covariates associated with the original epidemic trajectory.

To complement the visual comparisons, approximate normalized fit metrics were estimated from graphically extracted trajectories of the fitted curves. Because the target trajectories differ substantially in scale, the errors were normalized by the peak value of each target series. The resulting values were RMSE/peak ≈ 0.046 and MAE/peak ≈ 0.037 for FDH, and RMSE/peak ≈ 0.035 and MAE/peak ≈ 0.026 for FSH. These approximate metrics are consistent with substantial agreement between the fitted and target trajectories in both synthetic-target cases.

### 3.4 Variation in fitted parameters across conditions

Comparison of the fitted parameter sets across the four scenarios shows that the magnitude of parameter variation is not uniform. The most pronounced changes are observed in *p, ω*_*h*_, and *ω*_*w*_, whereas *d*_0_and *c*_0_remain comparatively stable. In addition, the derived composite quantity *p*(*ω*_*c*_ + *ω*_*h*_ + *ω*_*w*_), shown in Table 1, remains of the same order of magnitude across the four cases despite substantial variation in some of the individual fitted values. This behaviour is consistent with compensatory trade-offs among the mobility-related weights and the overall transmission scaling parameter. Although these comparisons do not constitute a formal parameter-identifiability analysis, they provide additional evidence that substantially different fitted parameter combinations can accompany successful reproduction of the respective target trajectories.

## 4. Discussion

### 4.1 Implications for structural and counterfactual interpretation

The central result of this study is that suppression of the vaccination-related input did not cause a substantial deterioration in the fit to the reference epidemic trajectory after re-estimation of the remaining adjustable parameters. This finding does not imply that vaccination had no epidemiological effect, nor does it establish that the vaccination-related component of the model is formally non-identifiable. It demonstrates a narrower but important point: within the fitted model examined here, close agreement with the reference epidemic trajectory can be retained even when the vaccination-related contribution is not active.

This result is relevant to the interpretation of counterfactual simulations. A counterfactual estimate obtained by modifying a particular model component depends not only on the model’s ability to reproduce the observed trajectory, but also on the extent to which the available observations constrain the contribution assigned to that component. If parameter re-estimation can compensate for suppression of a component while preserving a similar fit, goodness-of-fit alone cannot establish that the fitted contribution of that component is uniquely supported by the observed trajectory. Consequently, the present analysis does not demonstrate that the vaccination counterfactual reported in the original study is incorrect; rather, it shows that reproduction of the observed epidemic trajectory alone is insufficient to validate the structural interpretation on which such a counterfactual calculation depends.

The synthetic-target experiments provide complementary evidence concerning the flexibility of the fitted model. The model reproduced the main temporal features of qualitatively different target trajectories through parameter adjustment while retaining the observed mobility and variant-prevalence inputs. These experiments do not directly address whether the observed data uniquely identify a particular model structure, and they should not be interpreted as a formal test of structural identifiability. Instead, they show that the model possesses substantial trajectory-level flexibility under refitting. Together with the vaccination-input perturbation and the variation observed in fitted parameters, this flexibility illustrates how a close trajectory-level fit can coexist with considerable freedom in the fitted representation.

This distinction is particularly important for phenomenological models used for interpretation or counterfactual analysis. Flexibility is not itself a defect: it may be necessary for a low-dimensional model to approximate complex epidemic dynamics. However, the evidential meaning of a good fit depends on the purpose for which the model is used. A fit that is adequate for reproducing an observed trajectory does not necessarily provide the same degree of support for mechanistic interpretation or for counterfactual changes to individual model components.

### 4.2 Scope and limitations

Several limitations define the scope of the present analysis. Most importantly, the original HER-SYS infection data used by Kayano et al. were not publicly available. We therefore reconstructed pseudo HER-SYS data from the published infection trajectory and used simplified assumptions for the age distribution. The resulting analysis is not an exact reanalysis of the original epidemiological dataset. Nevertheless, the reconstructed data reproduced the published model behaviour sufficiently closely for the perturbation experiments performed here, and alternative age-distribution assumptions produced only minor changes in the fitted trajectory and approximate normalized fit metrics. These sensitivity tests reduce, but do not eliminate, uncertainty associated with the reconstructed input data.

The quantitative fit metrics reported here are also approximate because they were calculated from trajectories extracted graphically from the published and reconstructed figures rather than from the original numerical time series. They are intended to supplement visual comparison and to indicate the scale of differences among the fitted cases, not to provide a formal statistical model comparison.

More fundamentally, the analyses presented here do not constitute a formal structural- or practical-identifiability analysis. Nor do they compare a systematically defined set of competing model structures using likelihood-based, information-theoretic, or Bayesian model-selection criteria. The vaccination-input perturbation examines one deliberately selected structural change, while the synthetic-target experiments examine trajectory-level flexibility in the original model formulation. Accordingly, the results should not be interpreted as establishing a general one-to-many mapping between epidemic data and model structure.

These limitations also constrain the generality of the conclusions. The present study examines one published phenomenological epidemic model, and the degree of flexibility or compensatory behaviour observed here cannot be assumed to apply quantitatively to other models. The broader methodological relevance lies instead in the concrete demonstration that such behaviour can occur in a data-informed model used for counterfactual inference, and that close reproduction of an observed trajectory should therefore be distinguished from independent validation of the structural interpretation assigned to fitted model components.

### 4.3 Methodological implications

The methodological concerns illustrated by this case study are closely related to established discussions of identifiability, parameter degeneracy, structural uncertainty, and model equifinality [4]-[7]. The contribution of the present analysis is therefore not the introduction of a new formal framework for addressing these problems. Rather, it provides a concrete computational example of how they can become relevant when a fitted phenomenological model is used for structural interpretation and counterfactual inference.

Simple perturbation and refitting experiments such as those used here can be informative as exploratory stress tests. They cannot replace formal identifiability analysis or principled comparison of competing model structures, but they can reveal cases in which apparently successful reproduction of an observed trajectory is maintained under substantively altered fitting conditions. Such findings can help identify situations in which stronger inferential claims would require additional evidence beyond goodness-of-fit.

For models intended primarily for forecasting or descriptive reproduction, trajectory-level predictive performance may be the principal criterion of interest. For models used to assign effects to particular covariates or to estimate counterfactual outcomes, however, additional questions arise concerning whether those contributions are sufficiently constrained by the available data. The appropriate validation strategy should therefore depend on the intended inferential use of the model. In this sense, reproducible perturbation tests are best viewed as accessible complements to more formal analyses, rather than as a general diagnostic framework in their own right.

## 5. Conclusion

Using a published phenomenological COVID-19 model as a computational case study, we examined how strongly goodness-of-fit to an epidemic trajectory constrains the structural interpretation used for counterfactual inference. After reproducing the published model behaviour with reconstructed input data, we found that suppressing the vaccination-related input and re-estimating the remaining parameters caused only a limited deterioration in the fit to the reference epidemic trajectory. Complementary synthetic-target experiments further showed substantial trajectory-level flexibility under parameter adjustment, and comparison of fitted parameter sets revealed behaviour consistent with compensatory parameter trade-offs.

These findings do not imply that vaccination had no effect on COVID-19 transmission, that the vaccination counterfactual reported in the original study was incorrect, or that the Kayano–Nishiura model is unusable. Nor do the analyses constitute a formal demonstration of structural non-identifiability. Rather, they show that, in this case, close agreement with the observed epidemic trajectory alone does not uniquely support the structural interpretation required for vaccination-related counterfactual inference.

More generally, the results illustrate the importance of distinguishing successful trajectory reproduction from validation of the structural interpretation assigned to fitted model components. Simple computational perturbation and refitting tests can provide useful empirical stress tests of this distinction, while remaining complementary to formal identifiability and model-comparison analyses.

## Data Availability

All data and scripts generated in this study, including the reconstructed substitute for File2.Hersys_infection.csv and ready-to-use input files for reproducing the computational analyses, are publicly available from Zenodo at doi:10.5281/zenodo.22007200. The original R implementation of the Kayano–Nishiura model is available separately from the repository cited in Ref. [3]. The development version of the present repository is available on GitHub.

## References

[1] Brauer F, Castillo-Chavez C, Feng Z. Mathematical Models in Epidemiology:Springer; 2019.

[2] Kayano T, Ko Y, Otani K, et al. Evaluating the COVID-19 vaccination program in Japan, 2021 using the counterfactual reproduction number: Ssci Rep. 2023; 13:17762. doi: 10.1038/s41598-023-44942-6

[3] https://zenodo.org/records/15244462 Dataset and code for “Evaluating the COVID-19 vaccination program in Japan, 2021 using the counterfactual reproduction number” in Scientific Reports 2023 Oct 18;13(1):17762. doi: 10.1038/s41598-023-44942-6

[4] Cunniffe N, Hamelin F, Iggidr A, Rapaport A, Sallet G. Observability, Identifiability and Epidemiology --A survey: arXiv:2011.12202: doi: 10.48550/arXiv.2011.12202

[5] Raue A, Kreutz C, Maiwald T, Bachmann J, Schilling M, Klingmüller U, Timmer J. Structural and practical identifiability analysis of partially observed dynamical models by exploiting the profile likelihood: Bioinformatics 2009;25(15):1923. doi: 10.1093/bioinformatics/btp358

[6] Chowell G. Fitting dynamic models to epidemic outbreaks with quantified uncertainty: A primer for parameter uncertainty, identifiability, and forecasts: Infect Dis Model. 2017;2(3):379. doi: 10.1016/j.idm.2017.08.001

[7] Williams TG, Guikema SD, Brown DG, Agrawal A. Assessing model equifinality for robust policy analysis in complex socio-environmental systems: Environ. Model. Softw. 2020;134:104831. doi: 10.1016/j.envsoft.2020.104831

[8] COVID-19 Community Mobility Reports [Internet]. Japan: Google; 2019. Available from: https://www.google.com/covid19/mobility/

